# Is the Official Development Assistance in Guinea-Bissau “emergency” or “indispensable”? Perceptions of key stakeholders in the healthcare sector

**DOI:** 10.1101/2024.12.03.24318428

**Authors:** Anaxore Casimiro, Réka Maulide Cane, Michel Jareski Andrade, Luís Varandas, Isabel Craveiro

## Abstract

Official Development Assistance (ODA) is a type of financial support for low and medium-income countries to promote economic development and well-being. This study explores the perceptions of key healthcare stakeholders on the role of ODA in the health sector in Guinea-Bissau, focusing on maternal and child health. This qualitative study, conducted in May 2022 and February 2024, utilized semi-structured interviews with ten national and international healthcare stakeholders. Participants included present and past Ministry of Public Health officials and global partners in Guinea-Bissau. An interview script was used, including questions about the role and effectiveness of ODA in development, challenges in implementing health programs, coordination mechanisms, alignment of assistance with countries’ strategic documents, and recommendations for future improvements. Thematic categorical analysis, identification of standards, themes, and emerging relationships were identified in the participants’ responses. Findings reveal as main topics: (1) Participants considered that foreign aid is seen as a “humanitarian” or “emergency” assistance that complements the government and has been indispensable for project implementation and improvement of health indicators (2) Stakeholders reported significant challenges, including inadequate alignment of donor agendas with national health strategies, insufficient coordination among actors, and limited government ownership. Political instability undermines projects’ sustainability and long-term impact (3). It is necessary to reinforce the management capacity of the Ministry of Health and ensure that the guiding tools are used. ODA has been crucial to avoid the collapse of the health sector in Guinea-Bissau. However, a more cohesive strategy is essential to enhance ODA effectiveness in Guinea-Bissau. Recommendations include strengthening government leadership, aligning donor programs with national health priorities, and fostering improved coordination among health actors. Increasing government ownership of ODA projects could improve sustainability. This research offers crucial insights for policymakers, emphasizing the balance between ODA and long-term health improvements in Guinea-Bissau.

## Introduction

Official Development Assistance (ODA) is a form of government aid designed to promote the economic development and welfare of low- and middle-income countries like Guinea-Bissau (OECD, 2024).

In 2023, the support provided by the Development Assistance Committee (DAC) of the Organisation for Economic Cooperation and Development (OECD) reached approximately 223.7 billion U.S. dollars globally, representing an increase of 1.8% compared to 2022, mainly due to humanitarian aid provided to Ukraine (OECD, 2024). Regarding the African continent, around 42 billion U.S. dollars were allocated in the same year, showing a slight increase (2%) compared to 2022 (OECD, 2024).

Guinea-Bissau, a small country in West Africa, has a history of political and institutional fragility since its independence from Portugal in 1974. It is one of the most politically unstable countries in the world and highly prone to coups, having recorded four coups and 17 attempted coups since its independence (World Bank, 2024). In 2023, the estimated population was 2,153,339 (Department of Economic and Social Affairs, Population Division, 2024). The country ranks 179th out of 193 countries in the 2023/2024 Human Development Report, with a Human Development Index of 0.483, an average life expectancy of 59.9 years, and a GDP per capita of 1,880 dollars (2017 PPP) (UNDP, 2024). Data from the Harmonized Household Living Conditions Survey 2018/19 and 2021/22 showed that poverty increased from 47.7% in 2018 (equivalent to around 0.802 million poor people) to 50.5% in 2021 (equivalent to more than 0.886 million poor people) (Sering Touray & Nalourgo Kanigui Yaya Yeo, 2024).

Between 2002 and 2018, Guinea-Bissau received approximately 2.3 billion U.S. dollars in ODA directed towards various sectors, including infrastructure and social services, economic sectors, production sectors, raw materials assistance, and humanitarian aid. During this period, the OECD estimates that 364 million U.S. dollars were directly allocated to the health sector (including health programs, population, and reproductive health), representing 16.4% of official development aid allocated to all sectors. Of this amount, 197.4 million U.S. dollars, equivalent to 8.9%, were directed explicitly towards activities related to reproductive, maternal, neonatal, and child health (OECD CRS database organized by the authors). The country has heavily relied on official development aid in this area, accounting for 21.17% and 52.93% of its current health expenditures (World Bank, 2024). Key donors include the Global Fund, the United States of America, and Portugal (OECD CRS database data organized by the authors).

Based on available data, ODA has been crucial for improving maternal and child health in Guinea-Bissau, with positive effects seen in hospital infrastructure investments, health professional training, and vaccination awareness campaigns (Baldursdóttir et al., 2018; World Bank, 2024).

In addition to North-South cooperation, Guinea-Bissau has also benefited from South-South cooperation. One example of this cooperation is Cuba, which has been an important political actor in South-South cooperation, exporting innovative technologies in biotechnology and health equipment, as well as providing human resources (mainly doctors and nurses) to health systems notoriously lacking qualified labor (Buss et al., 2010).

Internationally, the need for alignment between governments, funders, and civil society in developing and implementing programs has been emphasized. This issue was focused on the Paris Agreement and the Accra Agenda and its subsequent alignment with the Sustainable Development Goals (SDGs) (Baldursdóttir et al., 2018).

It is well documented that donor influence, or their ability to guide the decisions or priorities of national health policymakers, occurs when beneficiary countries substantially depend on external funding (Khan et al., 2017). On the other hand, several articles have highlighted the problems that can arise from donor dominance in health policy processes in low-and middle-income countries. These issues include overshadowing existing programs and priorities of beneficiary countries, neglecting the strengths and absorption capacities of national health systems, and the difficulty of sustaining progress after donor funding ends (Khan, 2014, 2018).

It is crucial to understand the experiences of policymakers in low- and middle-income countries regarding donor influence and the mechanisms that sustain the relationships between low-income countries’ institutions and donors to reinforce national ownership of health policies and promote broader good governance (Khan, 2018).

Based on interviews with national and international partners working in Guinea-Bissau, this study seeks to comprehend the diverse factors influencing and shaping the direction of ODA in the health sector, encompassing projects and programs related to maternal and child health and the health system.

Our conceptual framework delineates the key components necessary for the successful execution of ODA in the health sector, as illustrated in Figure 1.

**Figure 1.**
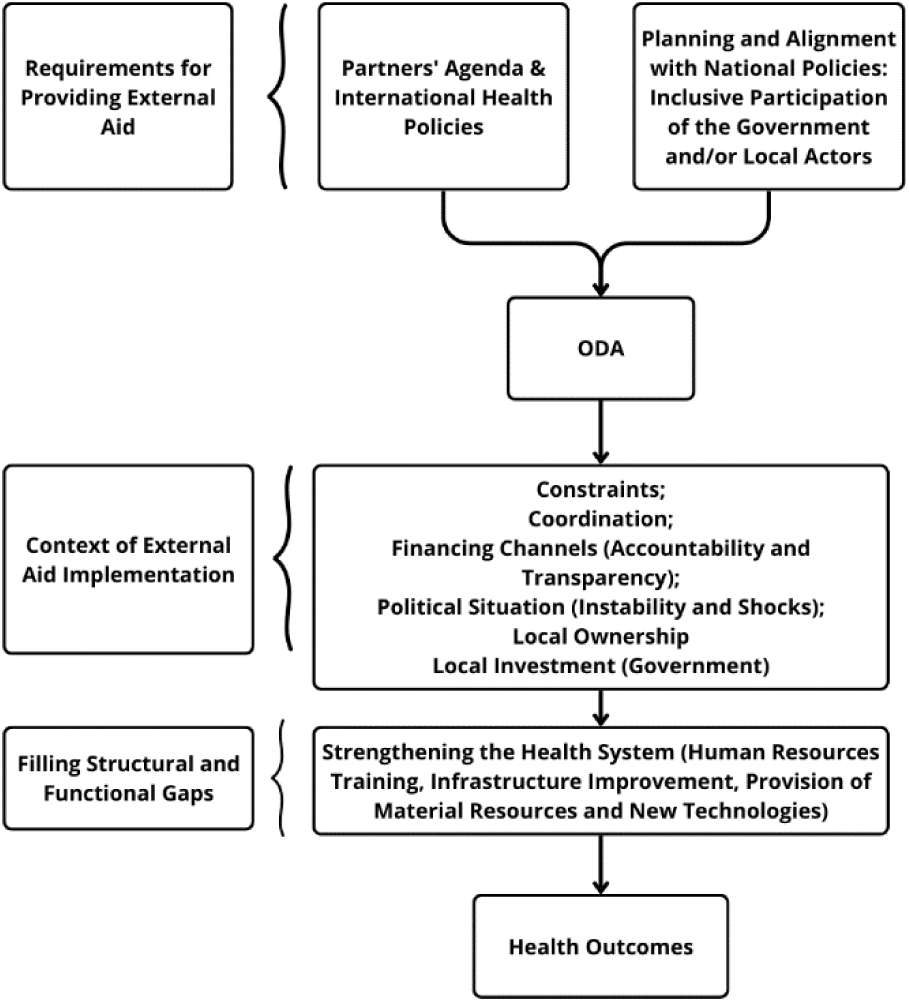
Conceptual framework on external development aid in the health sector. (adapted from Basar et al., 2020; Mitchell et al., 2020; Mataria, 2018).

For ODA to be effective, it is imperative to consider the following:

1. **The Agenda of Partners and International Health Policies** (Alesina & Dollar, 2000). The priorities established by international donors, global health organizations such as the World Health Organization (WHO), and Global Health Initiatives play a pivotal role in defining objectives and allocating resources, often giving precedence to vertical health initiatives (Hatefi & Allen, 2018; Kurihara & Moon, 2020).
2. **Planning and Alignment with National Policies**: The involvement of governments and local stakeholders in shaping programs and aligning them with national policies has been underscored in various initiatives over the years (Martini et al., 2012).
3. **The Context of ODA Implementation** The effectiveness of ODA is influenced by several factors in the local context, including:

a. Coordination: Effective coordination among the various actors involved in aid delivery is crucial to prevent duplication of efforts and ensure optimal use of resources (Bigsten & Tengstam, 2015; OECD, 2012).
b. Funding Channels: Transparency and accountability in funding mechanisms are essential for fostering trust and appropriate use of funds (Uzochukwu et al., 2018).
c. Political and institutional context: Political and institutional stability and crisis management capacity directly impact the funding and implementation of health programs (Alesina & Dollar, 2000).
d. Local Ownership: Active participation of the government and local communities in taking ownership of health programs is fundamental for the sustainability of initiatives (Brown, 2017; OECD, 2006).
e. Local Investment: The commitment of the local government to invest in health systems is vital for long-term sustainability, involving a shared vision of healthcare, a clear quality strategy, strong regulation, and continuous learning (Kruk et al., 2018).

## Materials and Methods

A qualitative research study was carried out in Guinea-Bissau between May 2022 and February 2024. Semi-structured interviews were conducted with key stakeholders involved in maternal and child health. Guidelines for ensuring rigor and reflexivity in qualitative research were followed, and the researchers completed the COREQ checklist for reporting qualitative data (Booth et al., 2014; Tong et al., 2007) attached as “S1 Appendix”.

### Participants and recruitment

Potential participants were purposively selected from key stakeholders at the central level of the national health service (particularly in maternal and child health) and global health and development cooperation partners. Selection criteria included health professionals with current or previous experience in the Ministry of Public Health of Guinea-Bissau, cooperation attachés, health program managers from various countries and international organizations operating in Guinea-Bissau, and representatives from non-governmental organizations (NGOs) working in the field of maternal and child health. Twenty key individuals were identified and invited to participate in the study. Ten participants (seven men and three women) accepted the invitation and were recruited for the study. Most participants had ten or more years of experience in the health or development cooperation field (Table 1). This research was approved by National Health Research Ethics Committee of Guinea-Bissau (n° 022/CNES/INASA/2021).

**Table 1.**
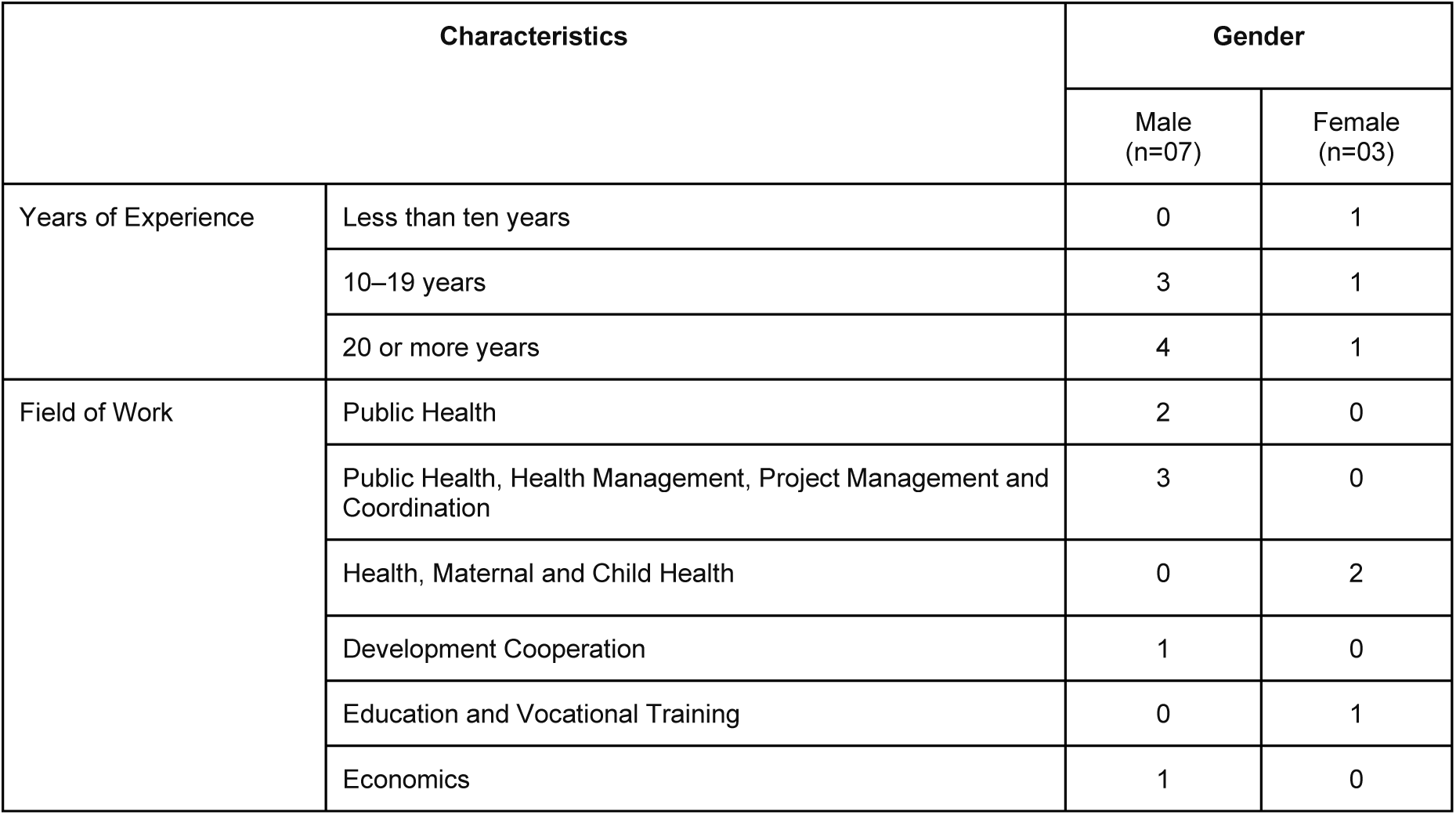
Characterization of the Interviewees.

### Data Collection

Key stakeholder’s in-depth interviews were conducted using a semi-structured topic guide developed by the study team. The guide (S2 Appendix) enabled a comprehensive exploration of the participants’ experiences and opinions on the effectiveness of official development assistance, challenges in implementing maternal and child health programs, coordination mechanisms, and suggestions for future improvements. Interviews were conducted by an experienced research team member (AC).

The interviews, conducted in Portuguese and English, lasted approximately 45 minutes each, and written informed consent was obtained from all participants. The interviews were recorded and transcribed verbatim using Notta.AI software (https://www.notta.ai/en).

### Analysis

Three researchers (AC, RC, MA) conducted data analysis using content analysis to identify patterns, themes, and emerging relationships in the participants’ responses. Content analysis is a research technique systematically used to analyse communication content to infer knowledge about the conditions of production and reception of these messages. It is widely applied in various fields, including social sciences, psychology, and healthcare, to study verbal and non-verbal communication (Bardin, 2004).

The transcriptions were entered into MAXQDA 24 software (MAXQDA, Software for qualitative data analysis, 1989 – 2024) to discern patterns and themes. A summary of key results, including relevant interview excerpts, was then created based on the identified thematic categories.

Ethical approval was obtained by National Committee for Ethics in Health Research, Ref. No. 022/CNES/INASA/2021. Informed written consent was obtained from all participants.

## Results

Ten participants, six global partners, and four local stakeholders were included in the present study.

Thematic analysis was done deductively and inductively. The themes emerging deductively reflected our analytical framework of the key components necessary for the successful execution of ODA in the health sector (Figure 1).

Inductively, focusing on a context like Guinea-Bissau, we propose classifying ODA into five categories: 1) The Role and Impact of ODA, 2) Constraints, Challenges, and Priorities of ODA in Health, 3) Donor Dynamics Over the Years, 4) Ownership of ODA in the Health Sector and Project Sustainability,5) Improving the Effectiveness of ODA in the Health Sector (Figure 2).

**Figure 2.**
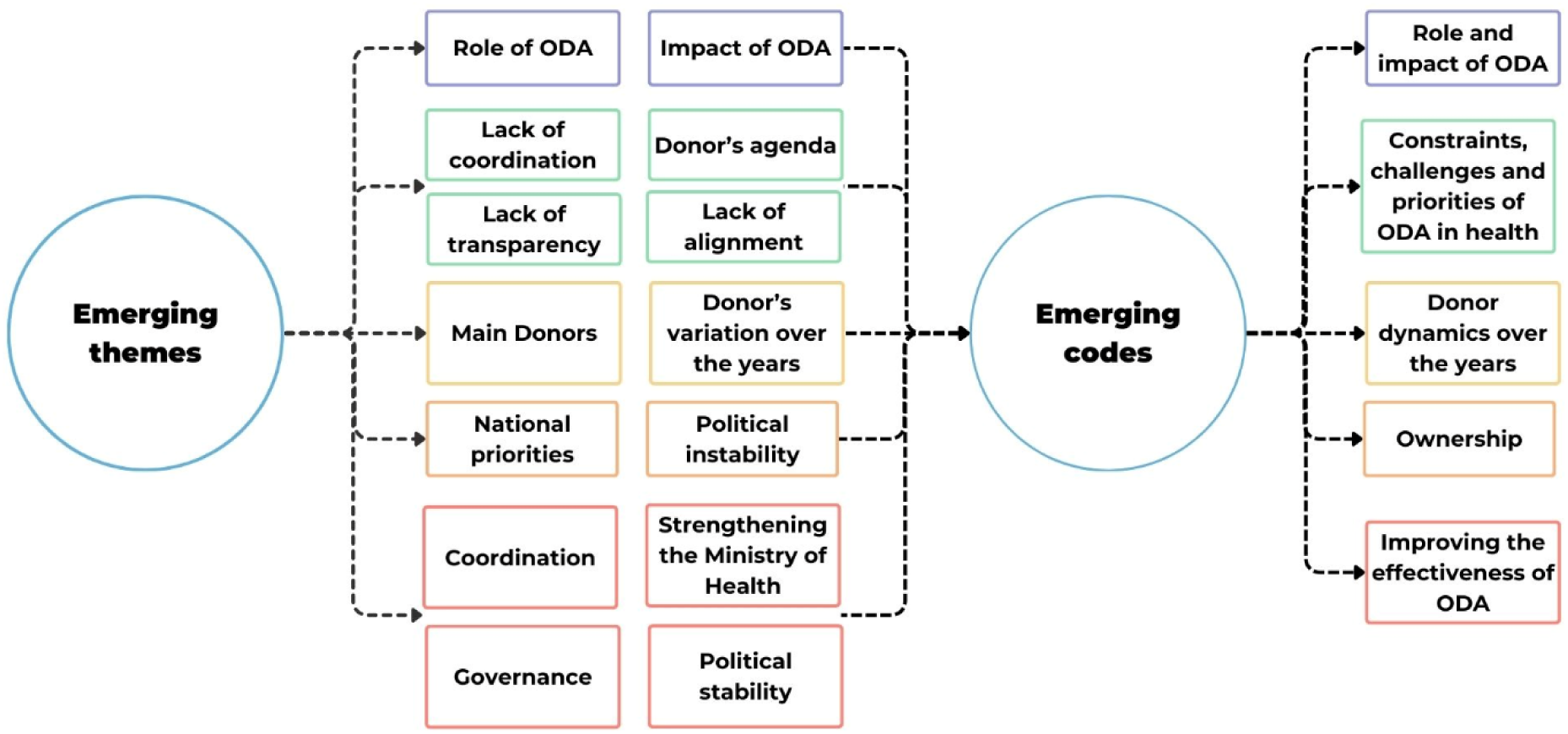
Main codes and themes emerging from the data deductively.

### The Role and Impact of Official Development Assistance (ODA)

When asked about their understanding of the role and impact of ODA, most interviewees (6/10) stated that ODA has been indispensable for implementing activities and projects in the social and health sectors. The mobilization of technical assistance has been fundamental in improving maternal and child health indicators and in preventing a catastrophe in areas such as tuberculosis, HIV/AIDS, or malaria. On the other hand, ODA is often viewed as “humanitarian” or “emergency” aid that complements the government rather than as aid for the country’s development, frequently not aligned with strategic goals but rather with the donors’ agendas.

> “The truth is that issues like tuberculosis, HIV/AIDS, malaria, and maternal and child health would have been catastrophic without these major actors funding them.” (Interviewee 008, Global Partner)

> “Essentially, we have somehow managed to hold on, and when I say ‘we,’ I mean the generality of international cooperation. At least those donors with greater financial muscle have clearly helped sustain the health indicators in this country. (…) It wouldn’t really be an aid for development in a stringent definition, but more like humanitarian or emergency aid, so to speak.” (Interviewee 008, male, Global Partner)

> “… a positive effect I can highlight is that the main donors, despite the successive crises, have never abandoned Guinea-Bissau, and this is a positive aspect.” (Interviewee 007, male, Global Partner)

Several interviewees (6 out of 10) noted that, in numerous instances, ODA constitutes the majority, or even the entirety, of the health sector budget. This external funding enables the introduction of innovations and supports previously identified projects that otherwise lacked the necessary resources for implementation. As a result, external aid often shapes and sometimes supplants the efforts and investments the Government of Guinea-Bissau should make in the social and health sectors. Nevertheless, data from 2000 to 2021 indicate that out-of-pocket expenditures have consistently accounted for the bulk of health investment in the country, placing a substantial burden on the population (World Bank, 2024).

> “… external aid, I think, has its negative sides, the consequences. It reduces the State’s effort to contribute to the health of women and children in the country…” (Interviewee 006, female, Global Partner)

> “External aid ends up constituting the majority, more than 90% of the budget, or in many cases, 100%, except in situations where the nature of the project itself requires a national counterpart.” (Interviewee 001, male, Local Stakeholder)

> “… Then you could say it’s essentially substituting government. This government is paying only salaries, and sometimes not on time, so yeah, government substitution, that’s what’s happening.” (Interviewee 010, male, Global Partner)

### Constraints, Challenges, and Priorities of ODA in Health

Interviewees mentioned several constraints, challenges, and priorities that affected the implementation of ODA in health.

#### Constraints

When asked about ODA’s constraints, the interviewees (5/10) criticized the lack of transparency in management, institutional instability, the lack of preparation among leaders, politicians, and managers, and donor-imposed agendas that do not align with the national agenda.

> “I specifically refer to when some projects, some aid packages, are already predetermined; the project to be implemented is already, therefore, preconfigured.” (Interviewee 001, male, Local Stakeholder)

> “All of this is very complicated in this country due to political instability.” (Interviewee 002, female, Global Partner)

#### Challenges

In the health sector, key challenges include poor coordination and inefficient resource management, insufficient implementation of strategic plans, heavy dependence on externally conditioned aid, limited investment in human resource capacity building, and political interference. Four out of ten interviewees highlighted the critical need for strong leadership at the government level to better align funding with national strategic policies, improve maternal and child health outcomes, combat corruption, and strengthen donor confidence in governmental structures. However, political instability and the frequent turnover of ministers have been significant barriers to the effective implementation of strategies over time.

> “…there must be policies where donors can also see leadership and look at Guinea-Bissau as a whole and perhaps redirect funding…” (Interviewee 006, female, Global Partner)

> “But if there were already a bit stronger capacity at the Ministry level, to plan health, to advocate for more resources in the state budget, to rationalize human health resources, to use the few resources available more correctly…” (Interviewee 002, female, Global Partner)

According to the interviewees, regarding the relationship between international donors and Guinean authorities, there is a concern among donors about the capacity of public institutions to “absorb” the principles of international cooperation and implement interventions jointly and collaboratively. On the other hand, the interviewees also criticized the lack of conditions for the Guinean government to contribute financially to health projects. Additionally, the importance of justifying the funds received (to the donors) and the need for dialogue with representatives of these funds to ensure that resources serve the country’s fundamental interests was highlighted. Other challenges mentioned by the interviewees include the lack of updated statistics in the health sector and poverty, which is seen as a factor that can lead to corruption and affect the performance of professionals.

> “…the truth is that there has been… some mistrust of donors in the capacity of public institutions to absorb what international cooperation can provide.” (Interviewee 008, male, Global Partner)

> “And the biggest drama, which is not only in Guinea-Bissau but in all African countries, is statistical problems. It’s like travelling without having rearview mirrors to see how things are (…), so performance does not escape corruption factors… Because real poverty is harsh and bitter…” (Interviewee 003, male, Global Partner)

#### Coordination Mechanisms for ODA in the Health Sector

When we delve deeper into the mechanisms for coordinating public development aid in this area, interviewees noted that the lack of coordination in the health sector in Guinea-Bissau has led to the duplication of interventions in certain areas and has hindered the intended impact of external aid. Efforts are underway to establish a coordination mechanism for external aid, but there is still a lack of communication and transparency among different actors.

> “The absence of a national-level coordination mechanism in the health sector, among others, has made it difficult to align the various supports.” (Interviewee 001, male, Local Stakeholder)

> “[…] there should be a hybrid coordination, meaning from the national side that receives and from the external side that gives.” (Interviewee 003, male, Local Stakeholder)

> “I think the greatest challenge is this. Not just to have a coordination effort, to know how much I’m going to put into certain areas but to present it directly to the State and have it accounted for in the General State Budget.” (Interviewee 005, male, Local Stakeholder)

Some interviewees mentioned the need to pressure the government for leadership and accountability in the health sector. Coordination should be hybrid, involving both the national side, which receives, and the external side, which gives. The lack of national coordination leads to certain regions receiving more funding than others. There is coordination in some programs, such as the vaccination program, but there is still little focus on effective coordination.

> “We need to pressure the government for leadership and accountability.” (Interviewee 002, female, Global Partner)

> “As for the partners, of course, as I was saying, they coordinate to avoid ‘overlap,’ but from the national side, the receiving side, there hasn’t been coordination. And what does this bring? Sometimes, we see situations where certain regions receive a ‘bomb’ of funding while others don’t.” (Interviewee 003, male, Global Partner)

> “If there isn’t strong leadership guiding all the partners’ funding, it ends up being disorganization in the health sector in particular.” (Interviewee 007, male, Global Partner)

#### Alignment of Funding with National Strategic Documents

When asked about the alignment of external development aid with national strategic documents, the interviewees (6/10) mentioned that there are strategic plans developed by the country that are considered by some donors in project development. However, these plans are often ignored after external funding is secured, and donors frequently work with their plans instead.

> “It is mandatory for us… we have to mention this. In one of our programs, it is quite evident that we have to make a reference to the national programming document.” (Interviewee 002, female, Global Partner))

> “Yes, yes…obviously. To begin with, partners do not finance on a casual basis. They finance based on an established plan, developed, discussed, and approved, and they finance based on a policy. They finance based on a national strategic plan and operational plans. Furthermore, they even go to the operational level. We are the ones who develop these plans, and we are the ones who elaborate them.” (Interviewee 003, male, Global Partner)

> “But, however, it is necessary to have the policy and have strategic plans to support the request for external funding. Once external funding is secured, it is almost as if the country’s strategic plan is forgotten.” (Interviewee 006, female, Global Partner))

The interviewees (7/10) also mentioned a lack of coordination and alignment among international partners in implementing health development projects in Guinea-Bissau. This results in projects not aligned with global and national strategies and the imposition of norms and methods that do not correspond to the country’s reality.

> “There is no coordination, nor alignment, nor capitalization of the projects.” (Interviewee 001, male, Local Stakeholder)

> “In my view, for all these plans, strategic documents of the Ministry of Health, both the National Health Development Plan and the Human Resources Plan, to be feasible and implemented, strong leadership and coordination of the health sector is needed.” (Interviewee 007, male, Global Partner)

#### Priorities

When asked about priorities for implementing ODA in the health sector, six out of ten interviewees highlighted vital issues such as enhancing coordination among various stakeholders, strengthening the leadership of the Ministry of Health, and improving both the stability and quality of human resources in the health sector, despite frequent changes in government.

> “… But we must have some non-politicized technical staff within the ministry, which do not change with each change of government. This is not acceptable. So, stop this spoiling system and depolitize it. This is a ministry that must have technicians. It is important…” (Interviewee 002, female, Global Partner)

> ”..Therefore, there is, and there must be policies that donors can also see the part of the leadership…” (Interviewee 006, female, Global Partner)

### Donor Dynamics Over the Years

Regarding the key actors in ODA in Guinea-Bissau, the majority of interviewees (6/10) highlighted that the donors supporting health in this country include the World Health Organization (WHO), the World Bank, the Global Alliance for Vaccines and Immunization (GAVI), the United Nations Children’s Fund (UNICEF), the United Nations Population Fund (UNFPA), the European Union (EU), non-governmental organizations (NGOs), and other institutions. The Integrated Program for the Reduction of Maternal and Child Mortality (PIMI, financed by the European Union) was important for maternal and child health in Guinea-Bissau.

> “The European Union, as you well know, has been working on maternal and child health since at least 2011…” (Interviewee 002, female, Global Partner)

> “In the case of the vaccination program, it’s GAVI…” (Interviewee 004, male, Local Stakeholder)

When asked about the variation of donors over the years, the interviewees (6/10) mentioned that they have been relatively constant in the health sector, with some bilateral partners coming and going. Civil society organizations, such as non-governmental organizations (NGOs), have received funding and have acted as implementing agencies. However, according to the interviewees, there is a perception that the funding volume has decreased recently. The most frequent donors include the United Nations, the European Union, the World Bank, the African Development Bank (AfDB), and the Global Alliance for Vaccination (GAVI). Germany is returning through the Economic Community of West African States (ECOWAS), while the United States has significantly reduced its bilateral aid package. Portuguese cooperation has also funded small projects. In the health sector, there is a stable framework of donors.

> “So, in the landscape of actors and funding, there hasn’t been a wide variation. There has been almost constancy, but we can notice that there is a decrease.” (Interviewee 003, male, Global Partner))

> “So far, it’s been the same; if it’s not the European Union, it’s the World Bank, the AfDB, the African Bank, they’ve been constant so far.” (Interviewee 007, male, Global Partner))

> “I would say that in the health sector, there has been some constancy…” (Interviewee 010, male, Global Partner)

### Ownership of ODA in the Health Sector and Project Sustainability

Some interviewees (4/10) addressed the importance of project ownership by the government and the lack of sustainability. They also emphasized that frequent changes in sector and program coordination constitute an obstacle to ownership and sustainability.

> “There is no ownership of projects and project results. Their impact on the system ends up failing.”. (Interviewee 001, male, Local Stakeholder)

> “And the infrastructures that could be created or improved with this funding end up deteriorating because there is no alignment or ownership. Because there are also changes in the coordination of sectors, the heads of sectors, programs, etc.” (Interviewee 001, male, Local Stakeholder)

> “It’s fashion, everyone says sustainability, it’s a requirement, but cannot sustain. When there’s no government engagement, it’s a joke.” (Interviewee 010, male, Global Partner)

Some participants (4/10) highlighted the need to empower the government to take project responsibility. However, there is concern about the government’s unwillingness to take ownership of the projects and justify the funds received. Additionally, interviewees (3/10) mentioned that the government does not oversee the governance structure, leading to parallel implementation. Political instability is also seen as an obstacle to project sustainability.

> “But if the government doesn’t have the will, meaning it’s not fulfilling its part, or we in the program aren’t fulfilling our part in justifying the funds and others. It’s hard for the government to take ownership of the funds.” (Interviewee 004, male, Local Stakeholder)

> “In terms of donors, we are considering mechanisms for the country to take ownership of projects, particularly in the maternal and child health area, but progress remains unchanged” (Interviewee 006, female, Global Partner)

### Recommendations for Improving the Effectiveness of ODA in the Health Sector

When questioned about possible mechanisms to improve the effectiveness of ODA in the health sector, most interviewees (6/10) recommended the importance of coordination and articulation among the different sectors involved in implementing health projects, including the private sector and civil society. They also emphasized the need for technical skills to identify funding sources and develop projects that meet the country’s needs, the need to review bureaucratic procedures to expedite fund acquisition and ensure good management, and the need to adopt a mindset focused on the usefulness of management tools and work at the roots to ensure the health system’s functioning as a whole.

> “[…] before starting project implementation, they should try to understand more or less, through the Ministry of Health, through Foreign Affairs, which they must go through, to understand the modalities in terms of coordination, the obligations of the institutions here in the country, what they should do in terms of accountability.” (Interviewee 001, male, Local Stakeholder)

> “It can’t be that every time the government changes, I understand, the minister, of course, has to work with trusted people who also express his policy, but we also need technicians, people who know how to manage, for example, the human resources in health, how to manage medicines, and all that.” (Interviewee 002, female, Global Partner)

> “What I believe can be improved is the bureaucratic procedure. […] And this usually delays the fulfilment of activities. I think this should be reviewed.” (Interviewee 004, male, Local Stakeholder)

Some participants (3/10) also highlighted the importance of coordination between funders and national institutions. They emphasized the need to separate health policy from politics and ensure that political changes do not impact technical staff. Donors should know the country’s needs and invest in sustainable projects.

> “It’s necessary […] to focus on the population, see the real difficulties of the population […]. By seeing this, you can summarize their needs and specifically know what to ask for, as well as increase the population’s education level and technicians who may respond.” (Interviewee 006, female, Global Partner)

> “Create structures within the Ministry of Health itself that can provide responses and political guidance in terms of this area of maternal health.” (Interviewee 007, male, Global Partner)

According to the interviewees (5/10), it is necessary to strengthen the Ministry of Health’s management capacity and ensure that guiding tools are used. Strategic, long-term thinking is needed to avoid political instability. Finally, the need for creating a “basket fund” as an aggregated funding source to implement a single program was also mentioned. Still, it was emphasized that the focus must be on the final objective and not be restricted to donors’ demands.

> “I think the creation, they can call it what they want, but creating management committees or coordination platforms for external aid.” (Interviewee 003, male, Global Partner)

> “So the idea is to have a single aggregated source of funding to implement a single program, a strategic plan.” (Interviewee 005, male, Local Stakeholder)

> “In a way, it’s also necessary to shield the public system. Shield it in the sense that, in a political change, in a political crisis, the change should be at the political level and not affect the technicians.” (Interviewee 006, female, Global Partner)

## Discussion

The present study aimed to analyze the role of ODA in the health sector, focusing on maternal and child health, from the perspective of key stakeholders in Guinea-Bissau. The results obtained indicate three main topics: (1) The participants in this study consider ODA to be “humanitarian” or “emergency” assistance that complements the government and has been indispensable for the implementation of activities and projects in the social and health sectors. It has been fundamental for improving maternal and child health indicators.; (2) the lack of coordination in the health sector has led to duplication of interventions in certain areas, limiting the intended impact of ODA. On the other hand, ODA is often not aligned with strategic objectives but rather with the specific goals of funders.; (3) it seems necessary to strengthen the management capacity of the Ministry of Health and ensure that guiding tools are utilized.

### ODA in Fragile Contexts: “Emergency” or “Indispensable”? How do priorities vary?

In contexts of conflict and political instability, health systems become extremely fragile, with negative consequences that persist for many years after the end of the crisis (Russo et al., 2017). Generally, development support organizations opt to withdraw from conflict regions, favouring countries with a greater capacity to manage aid. On the other hand, countries that do not “conform” to the donors’ single approach fall into disfavour, and their populations are doubly condemned to live in conditions of political instability and to be denied aid for the same reason (Russo et al., 2017). The interviewees highlighted the difficulties faced by Guinea-Bissau, marked by cyclical institutional crises, coups d’état, and frequent government changes, which impact the government’s role in fulfilling its duties, often limiting it to paying staff salaries. This reality affects its ability to attract and utilize external aid resources to improve health indicators, particularly maternal and child health, and strengthen the health system in the long term. These findings align with previous studies (Galjour et al., 2021), highlighting the challenges faced in Guinea-Bissau in establishing a stable political order post-independence, with pernicious effects on social sectors, particularly health. On the other hand, evidence indicates that a characteristic of countries and organizations that distribute ODA is shifting their priorities, mainly for geopolitical reasons. In Guinea-Bissau, historical examples include the total withdrawal of the Swedish International Development Authority - SIDA (1998) due to political instability (Einarsdóttir et al., 2005; Russo et al., 2017). Among the measures to mitigate the effects of the country’s political instability on the health sector, some authors (Galjour et al. 2021) recommend efforts focused on strengthening the capacities of health officials within the Ministry of Health of Guinea-Bissau to protect them from the effects of the country’s political instability. Other authors (Rasmussen et al., 2019) suggest establishing appropriate stock management systems and “backup” plans for periods of political instability to ensure that essential health system functions are maintained, thus increasing health system resilience. To achieve these recommendations, there must be flexible and participatory collaboration between the government sector and cooperation partners.

### Lack of Government Ownership of Projects and Related Aspects

The lack of government ownership of projects in Guinea-Bissau emerged as one of the central themes in the interviews. This issue makes donor-funded initiatives less effective. Local ownership is crucial for the success of development projects, as, without the engagement and commitment of the local government, projects often collapse once donors withdraw (Brautigam et al., 2004; Shumey, 2018; Easterly, 2008). According to the Paris Declaration, ownership is a *sine qua non* condition for applying the Principles of Aid Effectiveness. A development strategy with national ownership is a requirement for donor countries to align their aid with government priorities and the basis for harmonizing their development aid among themselves (OECD, 2006; Brown, 2017).

However, several interviewees linked the lack of ownership to the difficulty in leadership and assertiveness demonstrated by national authorities. The lack of leadership by national authorities has been widely described in the literature in various countries and contexts, some of them close to the reality of Guinea-Bissau, such as Mali and Ghana (Brown, 2017). Furthermore, political instability and the lack of government structure can undermine developing projects and their continuity after ceasing investments (Chauvet, 2010). On the other hand, the lack of sustainability and the flaws in investments were perceived by the participants in this study as a factor that creates financial dependency, hindering the development of sustainability and independence of the national government. These results corroborate evidence that there is an excessive focus on specific areas, such as maternal and child health, considered “trendy areas” (Moyo, 2009). Among the critical aspects to be improved for the effective use of all received and available resources are coordination and articulation between sectors—considered essential for enhancing ownership. Effectively coordinating efforts between the public, private, and civil society sectors can be crucial for maximizing the impact of aid (Buchner, 2011). In various contexts, there are sectoral coordination units for foreign assistance. The excess of project proposals from multiple donors is a challenge for sectoral coordination units, and much of development funding is not reflected in national budgets (Hill PS, Brown S, Haffeld, 2011).

Technical capacity building and funding identification are also crucial and should align with national plans to develop projects that meet local needs (Moss, 2006). However, for this to happen, bureaucratic procedures must be simplified. Political changes should not affect technical professionals or the implementation of strategies, particularly in maternal and child health when excessive administrative processes and the lack of skilled professionals can hinder effective development (Acemoglu, 2012; Birdsall, 2007; Azevedo, 2017), making it necessary to make more significant efforts to avoid this outcome.

A properly functioning health system must have the necessary ingredients, depending on financial, social, economic, environmental, and workforce resources. Although there is no “magic formula” to solve all the factors that make managing a health system in precarious contexts difficult, committed leadership—with a people-focused vision, especially for the most vulnerable—is one of the crucial factors for strengthening the national health system in Guinea-Bissau (Azevedo, 2017).

### Strengthening the Ministry of Health’s Management Capacity and Ensuring More Coordination and the Use of Guiding Tools

Most interviewees highlighted the need to strengthen the human resources of the Ministry of Public Health, which would improve management capacity, coordination, alignment with national priorities, and long-term project ownership. In this sense, the National Health Development Plan III-2023-2028 recognizes that a dedicated team for fundraising and diversification of funding sources should be created within the Ministry of Public Health, whose first mission will be to develop a medium-term Sector Financing Plan, which will be validated by the Health Sector Coordinating Committee (HSCC). This Sector Financing Plan is aligned with the NHDP and the entire strategy for the sector (Ministry of Public Health, National Health Development Plan III - 2023-2028).

The issue of partner activity coordination is recurring in the literature. It has been observed in other contexts, such as Kenya, where a study indicates that, although formal coordination structures exist, duplication, fragmentation, and misalignment of health system functions and stakeholder actions undermine sector coordination. These coordination challenges seem to be driven by weak governance, the effects of decentralization, and fragmentation caused by donor financing agreements (Nyawira L. et al., 2023).

Similar experiences in improving governance capacity in the health sector have been applied in Burundi, where the main contributions of these reforms were the separation of functions, transparency in management, and a meticulous description of administrative procedures (Peerenboom P. et al., 2014; Azevedo, 2017).

Some authors (Bazon, 2018) also suggest that it is necessary to ensure that public servants (including health professionals) are qualified (trained) and fairly compensated, providing more significant equity and justice and also limiting situations of corruption and abuse of power that can compromise the proper functioning of the health system.

### Recommendations for Improving the Effectiveness and Perception of Ownership of External Development Aid in the Health Sector and Project Sustainability

The lack of coordination in the health sector in Guinea-Bissau has led to the duplication of interventions and hampered the impact of external aid. Studies indicate that effective coordination is crucial for maximizing the benefits of international aid, avoiding redundancies, and ensuring the proper allocation of resources (Easterly, 2008). The absence of a well-defined coordination mechanism can result in fragmented efforts, where different entities repeat similar activities without a clear view of the overall needs of the sector (Bräutigam, 2004; Easterly, 2008).

The attempt to establish a mechanism for coordinating external aid is a positive step. Still, it faces significant challenges due to the lack of communication and transparency among the various actors involved. Inter-institutional coordination requires not only the creation of formal structures but also the development of a culture of cooperation and information sharing. (Lundsgaarde, 2010). Without these elements, coordination initiatives tend to be ineffective, perpetuating duplication problems and poor resource distribution (Moss, 2006).

To improve the effectiveness of coordination, the government must take a more proactive leadership role and be held accountable for its efforts in the health sector. Coordination should be hybrid, involving national and international donors, ensuring equitable distribution of resources and alignment of interventions with local needs (Shumey, 2018).

The lack of ownership of projects by the government of Guinea-Bissau has emerged as one of the central themes in interviews. This factor significantly undermines the impact and effectiveness of donor-funded initiatives. Local ownership is essential for the success of development projects, as without the involvement and commitment of the local government, projects tend to fail after donors withdraw (Brautigam et al., 2004; Shumey, 2018; Easterly, 2008). Moreover, political instability and lack of government structure can undermine ongoing projects and their continuity after investment cessation (Chauvet, 2010). On the other hand, the lack of sustainability and flaws in investments were perceived by study participants as a factor creating financial dependency of recipient countries, hindering the development of sustainability and national government independence. Significant aspects to be improved for effective use of all received and available resources include coordination and articulation among sectors—considered essential for enhancing ownership. Effective coordination of efforts between the public sector, private sector, and civil society can be crucial for maximizing the impact of aid (Buchner, 2011).

Technical capacity and identification of funding are also crucial and should be aligned with national plans, aiming to develop projects that address real local needs (Moss, 2006). However, for this, bureaucratic procedures must be simplified, and political changes should not affect technical professionals or the implementation of strategies, especially in maternal and child health. Excessive administrative processes and the lack of trained professionals can hinder effective development, necessitating additional efforts to avoid this outcome (Acemoglu, 2012; Birdsall, 2007).

## Conclusions

This study reveals that ODA has been vital for health financing in Guinea-Bissau, particularly for maternal and child health. However, our findings emphasize that it often functions more as “emergency” relief than a long-term development catalyst.

Through comprehensive data analysis and interviews, we identified several barriers that affect the effectiveness and influence of health policies and development programs. These include political and governmental instability, insufficient coordination among stakeholders, a misalignment between donor agendas and national health strategies, and weak leadership from local authorities.

To overcome these systemic challenges, fostering cohesive collaboration, aligning efforts with national priorities, and increasing the ownership and leadership of the Ministry of Public Health of Guinea-Bissau are essential. This shift would enable ODA to evolve from a short-term remedy into a transformative force for sustainable health development in the country.

This study offers valuable insights for policymakers, national stakeholders, donors, and other partners aiming to enhance the role and effectiveness of development assistance in fragile health contexts.

## Supporting information

**<u>S1 Appendix</u>.** <u>COREQ</u> (Consolidated Criteria for Reporting Qualitative Studies). (PDF).

**<u>S2 Appendix.</u>** Semi-Structured Interview Topic Guide (WORD)

## Data availability

The data supporting this study’s findings (e.g., interview timeline and interview justifying quotes) are available from the corresponding author [A.C] upon reasonable request.

## Data Availability

All relevant data are within the manuscript and its Supporting Information files.

## Acknowledgments

The authors acknowledge the contributions of the healthcare stakeholders who participated in this study.

